# Perfluoroalkyl Substances are increased in patients with Late-Onset Ulcerative colitis and induce Intestinal Barrier defects *ex vivo* in Murine Intestinal tissue

**DOI:** 10.1101/2020.06.03.20120550

**Authors:** Frida Fart, Samira Salihović, Aidan McGlinchey, Melanie G. Gareau, Matej Orešič, Jonas Halfvarson, Tuulia Hyötyläinen, Ida Schoultz

**Affiliations:** School of Medical Sciences, Örebro University, 702 81 Örebro, Sweden; School of Science and Technology, Örebro University, 702 82, Örebro, Sweden; Department of Anatomy, Physiology and Cell Biology, School of Veterinary Medicine, University of California Davis, Davis, CA, US; Turku Bioscience Centre, University of Turku and Åbo Akademi University, 20520 Turku, Finland; Department of Gastroenterology, Faculty of Medicine and Health, Örebro University, 702 81 Örebro, Sweden

**Author notes:** **Address for correspondence:** Ida Schoultz, PhD, School of Medical Sciences, Örebro University, Södra Grev Rosengatan 30, 703 30 Örebro university, Sweden. Shared senior authorship (Jonas Halfvarson, Tuulia Hyötyläinen, Ida Schoultz).

**Keywords:** PFAS, bile acids, inflammatory bowel disease

## Abstract

**Background:** Environmental factors are strongly implicated in late-onset of inflammatory bowel disease. Here, we investigate whether high exposure to perfluoroalkyl substances correlates with (1) late-onset inflammatory bowel disease, and (2) disturbances of the bile acid pool. We further explore the effect of the specific perfluoroalkyl substance perfluoronoctanoic acid on intestinal barrier function in murine tissue.

**Methods:** Serum levels of perfluoroalkyl substances and bile acids were assessed by ultra-performance liquid chromatography coupled to a triple-quadrupole mass spectrometer in matched samples from patients with ulcerative colitis (n=20) and Crohn’s disease (n=20) diagnosed at the age of ≥55 years. Age and sex-matched blood donors (n=20), were used as healthy controls. *Ex vivo* Ussing chamber experiments were performed to assess the effect of perfluoronoctanoic acid on ileal and colonic murine tissue (n=9).

**Results:** The total amount of perfluoroalkyl substances was significantly increased in patients with ulcerative colitis compared to healthy controls and patients with Crohn’s disease (*p*<0.05). *Ex vivo* exposure to perfluoronoctanoic acid induced a significanlty altered ileal and colonic barrier function. The distribution of bile acids, as well as the correlation pattern between (1) perfluoroalkyl substances and (2) bile acids, differed between patient and control groups.

**Conclusion:** Our results demonstrate that perfluoroalkyl substances levels are increased in patients with late-onset ulcerative colitis and may contribute to the disease by inducing a dysfunctional intestinal barrier.

## Introduction

Inflammatory bowel disease (IBD), including the two main subtypes Crohn’s disease (CD) and ulcerative colitis (UC), is a chronic immune-mediated disease of the gastrointestinal tract. The burden of IBD is rising globally, and especially elderly-onset disease is becoming increasingly common [1,2].

The disease is thought to develop as a result of complex interactions between environmental factors, genetic susceptibility and altered gut microbiota. The gut mucosal barrier maintains homeostasis and prevents the entry of pathogenic bacteria and other harmful substances whilst simultaneously absorbing essential nutrients [3]. Some environmental risk factors, such as emulsifiers and heavy metals, may in fact mediate their effects *via* modulation of the mucosal barrier [4,5]. One of the main chemical groups that humans are exposed to through the diet is perfluoroalkyl substances (PFAS), which are human-made chemicals with a long biological half-life in humans. PFAS have been extensively used since the 1950s, in non-stick cookware and as flame retardants, amongst other uses. Even though the use of some PFAS such as perfluorooctane sulfonate (PFOS) and perfluoronoctanoic acid (PFOA) have been either prohibited or restricted, exposure to these substances still occurs as they as they, due to their long half-life, remain in the environment. Moreover, several of the current substitutes within the PFAS substance class, such as fluorotelomer alcohols, are precursors of several toxic PFAS, including PFOA [6]. An increased incidence of UC has previously been reported in response to high exposure to PFOA through contaminated drinking water in mid-Ohio, United States (U.S) [7]. This observation is supported by findings from animal studies, where PFOA has been reported to alter the immune response and the barrier function *via* regulation of various cytokines and tight junction proteins [8].

Recently, it has also been shown that PFAS enter the same enterohepatic circulation as bile acids (BAs) and that PFAS may impact the absorption of these BAs in the small intestine [9,10]. The BAs are synthesized from cholesterol in the liver, and these primary BAs are then conjugated either with glycine or taurine, and transported through the biliary system into the small intestine where they can be modified to become secondary BAs by the gut microbiota. Beyond facilitating the digestion and absorption of lipids in the small intestine, BAs possess antibacterial properties, of relevance for the gut mucosal defence, including intestinal permeability [9]. Dysregulation of intestinal BAs has further been implicated in the pathogenesis of IBD [11,12] and *ex vivo* exposure to BAs have been shown to induce an increase in intestinal permeability and bacterial uptake across the colonic mucosa [13]. Current knowledge suggests that late-onset IBD is more linked to environmental risk factors and is associated with a lower genetic risk score, as compared to earlier onset [14]. We, therefore, hypothesised that chronic exposure to PFAS could contribute to the development of late-onset IBD, by altering the intestinal barrier function or indirectly by interfering with the BAs metabolism and, thereby alter the intestinal barrier function. Here we investigate levels of PFAS and their relation to primary and secondary BAs in patients who developed IBD later in life. Moreover, we explore the effect of PFOA, one of the major PFAS, on the intestinal barrier function, by performing *ex vivo* experiments of ileal and colonic murine tissue using the Ussing Chamber.

## Material and Methods

### Study design

This is a case-control study where serum concentrations of PFAS and BAs were assessed in patients with late-onset IBD, defined as UC or CD diagnosed ≥55 years (cases), and age- and sex-matched blood donors, without any history of chronic gastrointestinal disease, i.e. healthy controls (HC). Comparisons with late-onset CD patients were also stratified for disease location, since BAs are reabsorbed in the terminal ileum. Based on the observed association between exposure to PFOA and late-onset disease, we investigated the effect of PFOA on ileal and colonic barrier function in wild type mice using the Ussing Chamber methodology.

### Study population

The samples from IBD patients used in this study were obtained from a previously described cohort [15]. Briefly, patients with CD or UC were consecutively recruited at the outpatient IBD clinic of Örebro University Hospital, Sweden. Blood samples were collected after receiving written informed consent and serum was separated according to standard operating procedures [16], aliquots were stored at −80°C until further analysis. A diagnosis of IBD was based on internationally accepted clinical, endoscopic, radiologic and histologic criteria [17]. The Montreal classification was used for assessment of disease characteristics [18]. A sample set of 20 late-onset UC and 20 late-onset CD patients, whom was diagnosed at age ≥55, matched by age (± 5 years), sex and disease duration (± 5 years) were selected. HCs were matched by age (± 5 years) and sex (median age 59 years, IQR 56-61 years). Clinical and demographic characteristics of IBD patients included in the study are shown in Table 1. None of the patients were included at disease onset, and blood samples were obtained, at a median of 7 to 8 years after diagnosis.

**Table 1.**
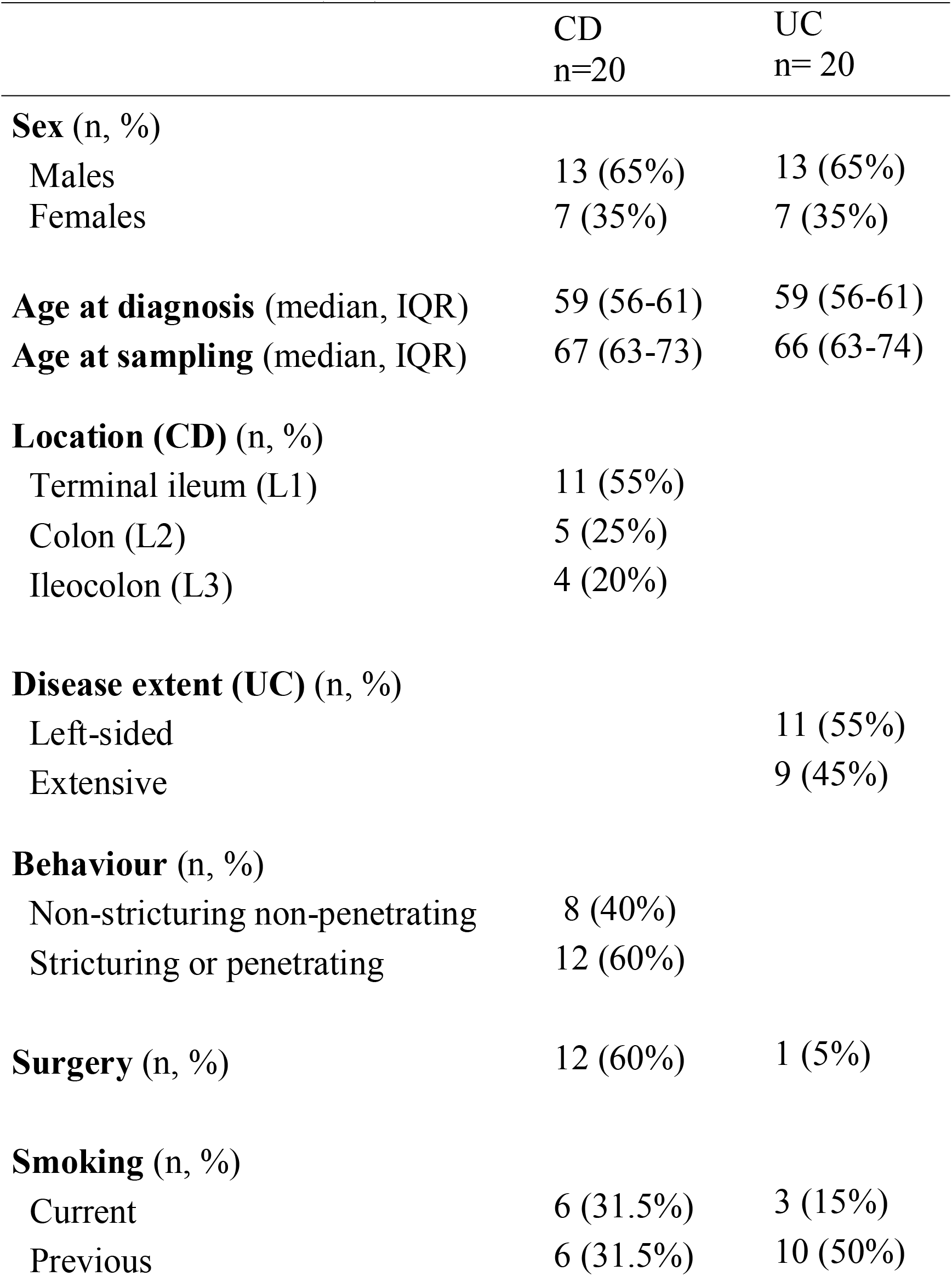
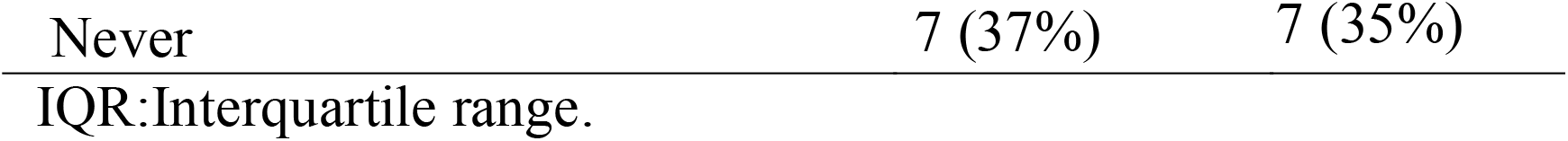
Clinical characteristics of patients with Crohn’s disease (CD) and ulcerative colitis (UC) CD UC

|  | CD<br>n=20 | UC<br>n= 20 |
| --- | --- | --- |
| **Sex (n, %)** |  |  |
| Males | 13 (65%) | 13 (65%) |
| Females | 7 (35%) | 7 (35%) |
| **Age at diagnosis (median, IQR)** | 59 (56-61) | 59 (56-61) |
| **Age at sampling (median, IQR)** | 67 (63-73) | 66 (63-74) |
| **Location (CD) (n, %)** |  |  |
| Terminal ileum (L1) | 11 (55%) |  |
| Colon (L2) | 5 (25%) |  |
| Ileocolon (L3) | 4 (20%) |  |
| **Disease extent (UC) (n, %)** |  |  |
| Left-sided |  | 11 (55%) |
| Extensive |  | 9 (45%) |
| **Behaviour (n, %)** |  |  |
| Non-stricturing non-penetrating | 8 (40%) |  |
| Stricturing or penetrating | 12 (60%) |  |
| **Surgery (n, %)** | 12 (60%) | 1 (5%) |
| **Smoking (n, %)** |  |  |
| Current | 6 (31.5%) | 3 (15%) |
| Previous | 6 (31.5%) | 10 (50%) |

IQR: Interquartile range.

### Analysis of BAs and PFAS

The levels of PFAS and BAs were assessed by ultra-performance liquid chromatography coupled to a triple quadrupole mass spectrometer as previously described [19]. Briefly, all samples were prepared and analysed simultaneously in a randomised order in one batch to minimise potential bias. A positive and negative control, as well as certified reference serum (NIST SRM 1957) were included in the analyses to assess accuracy and consistency.

### Ex vivo Ussing Chamber experiments

The Ussing chamber experiments were performed as described previously [20]. Briefly, segments of distal ileum and proximal colon from adult (6-8 week, n=9) wild type mice (C57BL/6, Jackson Labs, bred in-house at UC Davis, CA, US) were excised. The segments were cut along the mesenteric border and mounted in Ussing chambers (Physiologic Instruments, San Diego, CA, US), exposing 0.1 cm^2^ of tissue area with circulating oxygenated Ringer’s buffer (4 ml) maintained at 37°C. Additionally, glucose (10 mM) was added to the serosal buffer as a source of energy, which was balanced osmotically by mannitol (10 mM) in the mucosal buffer. Agar-salt bridges were used to monitor potential differences across the tissue and to inject the required short□circuit current (Isc) to maintain the potential difference at zero, as registered by an automated voltage clamp. A computer connected to the voltage clamp system recorded Isc and voltage continuously, and the data were analysed using acquisition software (Acquire and Analyze; Physiologic Instruments, San Diego, CA, US). Baseline Isc values (μA/cm^2^), indicative of active ion transport, were obtained at equilibrium, approximately 15 min after the tissues were mounted. PFOA (Sigma-Aldrich, San Francisco, CA, US) was added to both the serosal and mucosal side of the chamber in the concentrations of 100 μM and 10 μM at baseline. Conductance (G; mS/cm^2^) was used to assess tight junction permeability and mucosal to serosal flux of 4KDa FITC-labeled dextran (Sigma-Aldrich) over time (sampled every 30 minutes for 2 hours) was used to assess macromolecular permeability. After completion of the FITC flux measurements, tissues were treated with carbachol (CCh, 10^−5^ μM; Sigma-Aldrich) to assess the stimulated secretory response to PFOA.

### Statistical considerations

For statistical analyses of PFAS and BA concentrations, missing data/zeroes were replaced with imputed values calculated as half the minimum nonzero value for that substance, log2 transformed and scaled to zero mean and unit variance. The statistical analyses were performed using IBM SPSS statistics, MetaboAnalyst 4.0[21] and MetScape 3 for CytoScape.[22] In SPSS, a general linear model was fitted to investigate the associations between PFAS and BAs across the groups, after controlling for confounding factors (age and the year of sampling). To show the direction of the change between groups, the data is presented as fold change (*i*.*e*. ratio) between CD, UC and HC. Correlation analyses were done using the Spearmann correlation analyses. The Debiased Sparse Partial Correlation algorithm was used for estimating partial correlation networks, as visualised by the MetScape4 version with cut-off values of correlations between ± 0.22 to 0.75. When stratifying for ileal and colonic disease location in CD patients, Mann-Whitney U analysis was performed. For analysis of Ussing Chamber experiments Kruskal-Wallis and the Mann-Whitney U-analysis (GraphPad Prism 8; San Diego, CA, US) was used to assess intestinal permeability, FITC-dextran passage and stimulated secretory response. Values of *p*<0.05 were considered to be statistically significant. The degree of significance for Ussing Chamber experiments were designated as follows: *p*<0.05 [*], *p*<0.01 [**], non-significant [ns].

### Ethical considerations

The Local Ethics Committee approved the study (Dnr: 2006/245), and all patients, as well as, HCs gave their written informed consent. All animal procedures and protocols were approved by UC Davis animal care and use committee (IACUC #2007)

## Results

### Serum concentrations of PFAS in late-onset UC and CD

The concentrations of PFAS were assessed in serum samples (Table S1). The total level of PFAS (∑PFAS, *p*=0.03), PFOS (*p*=0.03) and PFOA (*p*=0.02) were found to be significantly increased in UC as compared to HC (Table 2). In contrast, the PFAS levels were not significantly different in CD and HC. Similarly, UC had significantly higher levels of ∑PFAS (*p*=0.03), PFOS (*p*=0.03) perfluorononanoic acid (PFNA, *p*=0.03), perfluorodecanoic acid (PFDA, *p*=0.005) and perfluoroundecanoic acid (PFUnDA, *p*=0.03), as compared to CD. A correlation network analysis further showed that disease is the major factor influencing the serum level of PFOA and that PFAS were closely associated with the BAs (Figure 1).

**Table 2.**
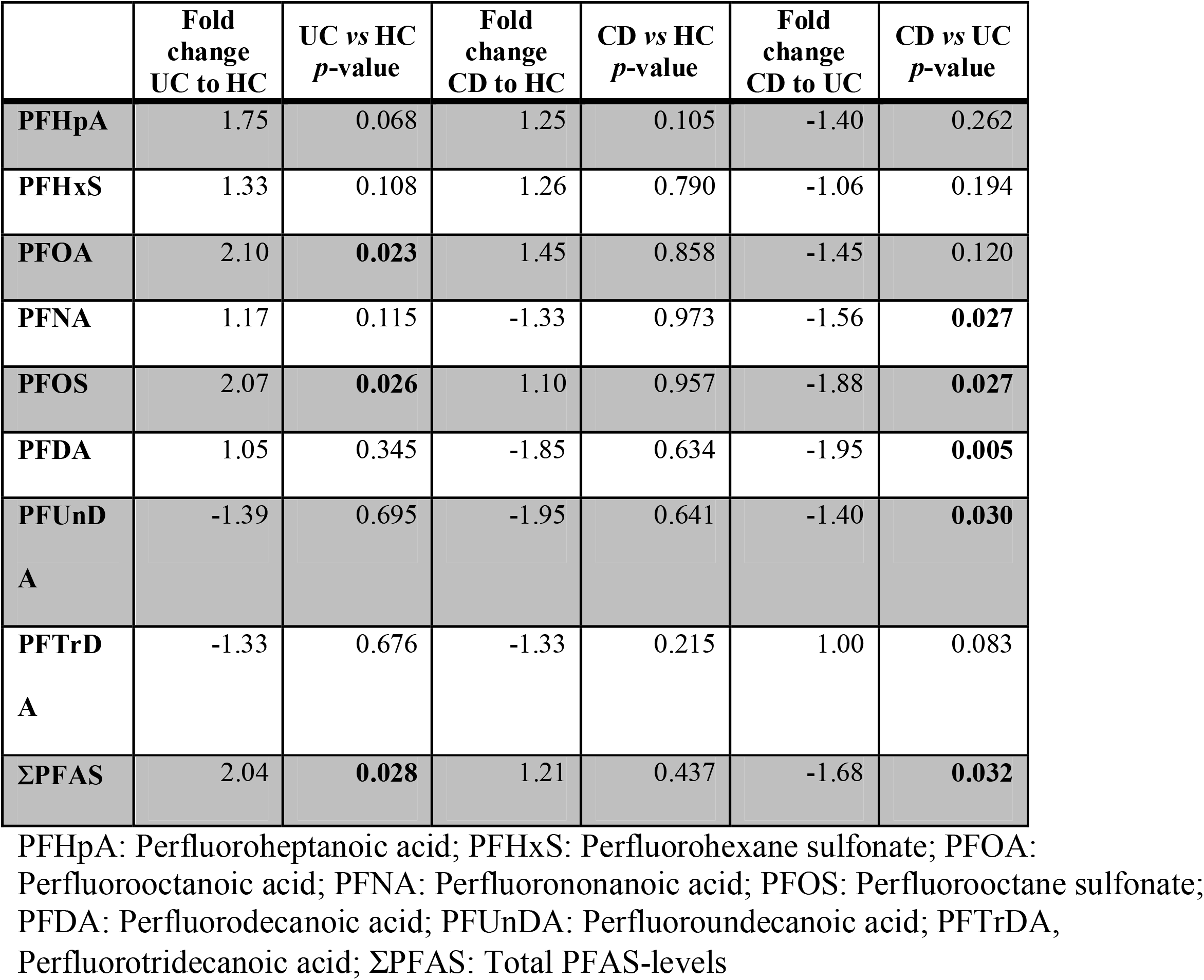
Comparisons of fold change ratio of serum perfluoroalkyl substances (PFAS) concentration in ulcerative colitis (UC), Crohn’s disease (CD) and healthy controls (HC) with adjusted *p*-values. A negative fold change value represents a decrease in PFAS-levels. Significantly changing PFAS (*p*<0.05) are marked in bold.

**Figure 1.**
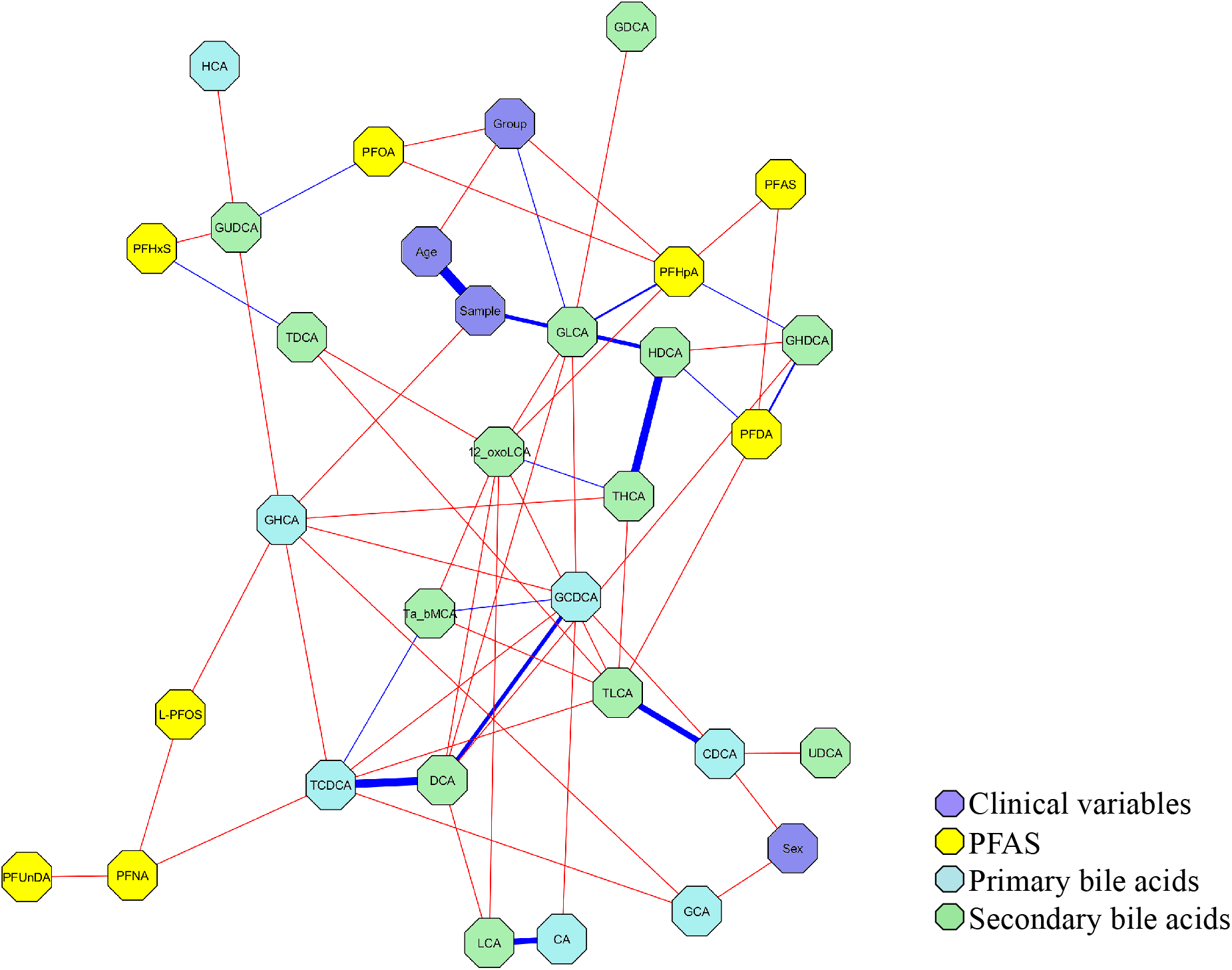
Partial correlation network projection of (1) clinical variables group (UC, CD and HC), age and sex, (2) PFAS, (3) primary BAs and (4) secondary BAs. Edge colors: blue for negative correlations and red for positive correlations, the thickness of the line shows the strength of the correlation. Edge ranges adjusted between ± 0.22 to 0.75.

### Ussing Chamber experiments

Intestinal permeability was assessed by measuring G for tight junction permeability and flux of FITC-dextran as an indicator of macromolecular permeability. *Ex vivo* exposure of intestinal murine tissue to PFOA at 100 μM resulted in a significant increase in ileal permeability at 60 minutes compared to baseline. An increased colonic permeability was observed at 60 minutes after exposure to 10 and 100 μM PFOA compared to baseline. A similar pattern was observed in vehicle (Figure 2a-b). Exposure to 10 μM PFOA induced an increased flux of FITC-dextran across ileal tissue (p=0.02). A trend was observed in colonic tissue but was not statistically significant (p=0.06) (Figure 2c-d). Stimulated ion transport across the ileal tissue, assessed by ΔIsc, was not impaired after exposure to PFOA. However, an increased response to CCh after exposure to 10 μM and 100 μM PFOA was observed across colonic epithelium compared to vehicle (DMSO) (Figure 2 e-f).

**Figure 2.**
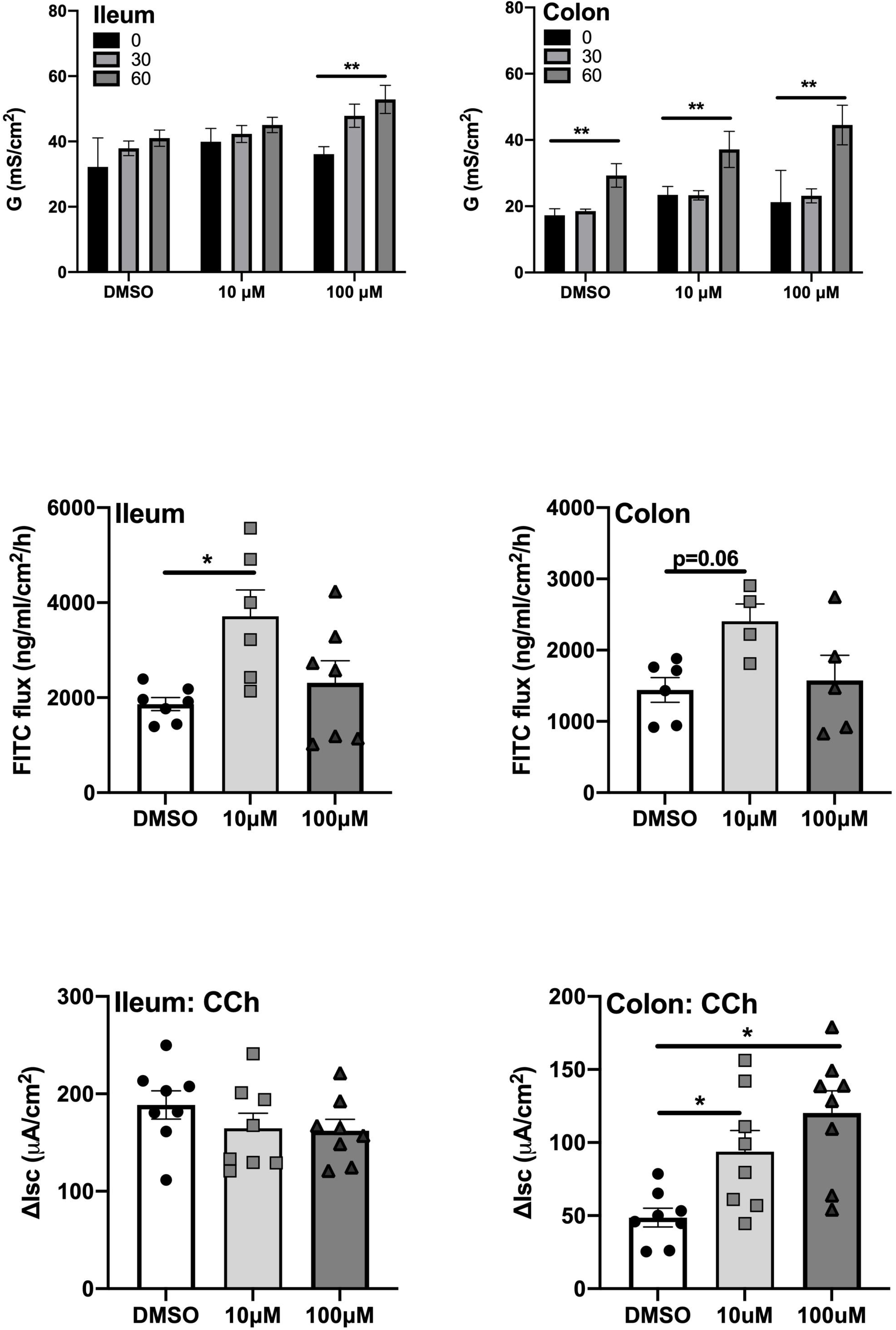
The effect of *ex vivo* exposure to PFOA on intestinal barrier function as measured by conductance, A) ileal and B) colonic conductance (G), macromolecular passage as assessed by FITC-dextran flux across C) ileal mucosa and D) colonic mucosa and stimulated secretory response to Charbachol, E) ileal and F) colonic short circuit current (Isc) after exposure to 10 μM, 100 μM PFOA or vehicle (DMSO). N=7-9. Data are presented as median with range (min/max), significance levels are calculated with Kurskal-Wallis and Mann-Whitney U-analysis, (*p*<0.05 [*], *p*<0.01 [**], non-significant [ns]).

### Serum BAs concentrations in UC and CD patients

The concentrations of BAs were assessed in serum samples (Table S1). The distribution of the BAs in the different groups are illustrated in Figure 3. The levels of primary BAs (free and conjugated BAs) were found to be decreased in UC as compared to HC (*p*=0.02), as shown in Table 3. At the same time, no differences were observed in secondary BAs or the total level of taurine-conjugated BAs taurine-conjugated BAs (ΣTBA) and glycine-conjugated BAs (ΣGBA). Primary BAs were decreased in CD as compared to HC (*p*=0.02) and UC (*p*=0.0001), except for increased levels of free chenodeoxycholic acid (CDCA) in CD. Moreover, the ratio of the two primary BAs, cholic acid (CA) and CDCA (CA/CDCA) was decreased in CD *vs* HC and UC (*p*=0.02), while no difference was observed between UC and HC. The total level of TBA was decreased in CD *vs*. HC (*p*=0.003), while no differences were found in the total level of GBA. The secondary conjugated BAs were found to be increased in CD patients as compared to HC (*p*=0.007).

**Table 3.** Comparisons of fold change ratio of serum bile acids (BAs) concentration in ulcerative colitis (UC), Crohn’s disease (CD) and healthy controls (HC) with adjusted *p*-values. A negative fold change value represents a decrease in BA levels. Significantly changing BA (*p*<0.05) are marked in bold.

| BA | Type of BA | Fold change<br>UC to HC | UC vs HC<br>*p*-value | Fold change<br>CD to HC | CD vs HC<br>*p*-value | Fold change<br>CD to UC | CD vs UC<br>*p*-value |
| --- | --- | --- | --- | --- | --- | --- | --- |
| CA | primary (free) | -1.21 | 0.0839 | 1.25 | 0.4587 | 1.51 | **0.0017** |
| CDCA | primary (free) | -1.10 | **0.0100** | 2.54 | **0.0056** | 2.78 | **0.0001** |
| TCDCA | primary<br>taurine-conjugated | -1.65 | 0.1844 | -7.03 | **0.0022** | -4.26 | 0.0804 |
| GCA | primary<br>glycine-conjugated | -1.88 | 0.0522 | -2.90 | **0.0133** | -1.54 | **0.0482** |
| GHDCA | primary<br>glycine-conjugated | -3.41 | 0.4722 | -1.69 | 0.8270 | 2.02 | 0.1238 |
| GCDCA | primary<br>glycine-conjugated | -1.49 | **0.0354** | -1.87 | 0.1380 | -1.26 | 0.0443 |
| Tα+βMCA | secondary<br>taurine-conjugated | -2.05 | **0.0010** | -9.85 | **0.0519** | -4.80 | 0.2440 |
| THCA | secondary<br>taurine-conjugated | -7.62 | 0.4676 | -160.00 | **0.0022** | -21.00 | 0.1658 |
| TDCA | secondary<br>taurine-conjugated | -1.12 | 0.9974 | -2.32 | 0.1070 | -2.08 | 0.1123 |
| TLCA | secondary<br>taurine-conjugated | -1.09 | 0.0696 | -3.43 | **0.0082** | -3.14 | **0.0225** |
| GHCA | secondary<br>glycine-conjugated | -1.99 | 0.1279 | -2.77 | **0.0002** | -1.40 | 0.1297 |
| GUDCA | secondary<br>glycine-conjugated | -1.63 | 0.1768 | -1.29 | 0.1653 | 1.26 | 0.0668 |
| GDCA | secondary<br>glycine-conjugated | -1.81 | 0.1511 | -2.32 | 0.2764 | -1.28 | 0.2759 |
| GLCA | secondary<br>glycine-conjugated | -3.10 | **0.0238** | -2.61 | **0.0293** | 1.19 | 0.3195 |
| UDCA | secondary | 1.25 | 0.6504 | 2.89 | **0.0047** | 2.30 | **0.0340** |
| HDCA | secondary | 1.68 | 0.4317 | 4.10 | **0.0016** | 2.44 | 0.0088 |
| 12-oxoLCA | secondary | 1.22 | 0.2645 | -1.59 | **0.0480** | -1.94 | 0.1143 |
| DCA | secondary | -1.35 | 0.1143 | 1.43 | 0.5410 | 1.93 | 0.3599 |
| LCA | secondary | 1.45 | 0.7307 | -1.25 | 0.8851 | -1.81 | 0.5002 |
| ΣPrimary<br>BA |  | -1.67 | **0.0173** | -2.05 | **0.0223** | -1.23 | **0.0001** |
| ΣSecondary<br>BA |  | -1.35 | 0.5424 | 1.59 | **0.0066** | 2.14 | 0.0646 |
| ΣTaurine<br>conjugated<br>BA |  | 3.37 | 0.3658 | -4.72 | **0.0024** | -15.89 | 0.0945 |
| **ΣGlycine**<br>**conjugated**<br>**BA** |  | -1.59 | 0.1192 | -10.26 | 0.0886 | -6.45 | 0.1227 |

**Figure 3.**
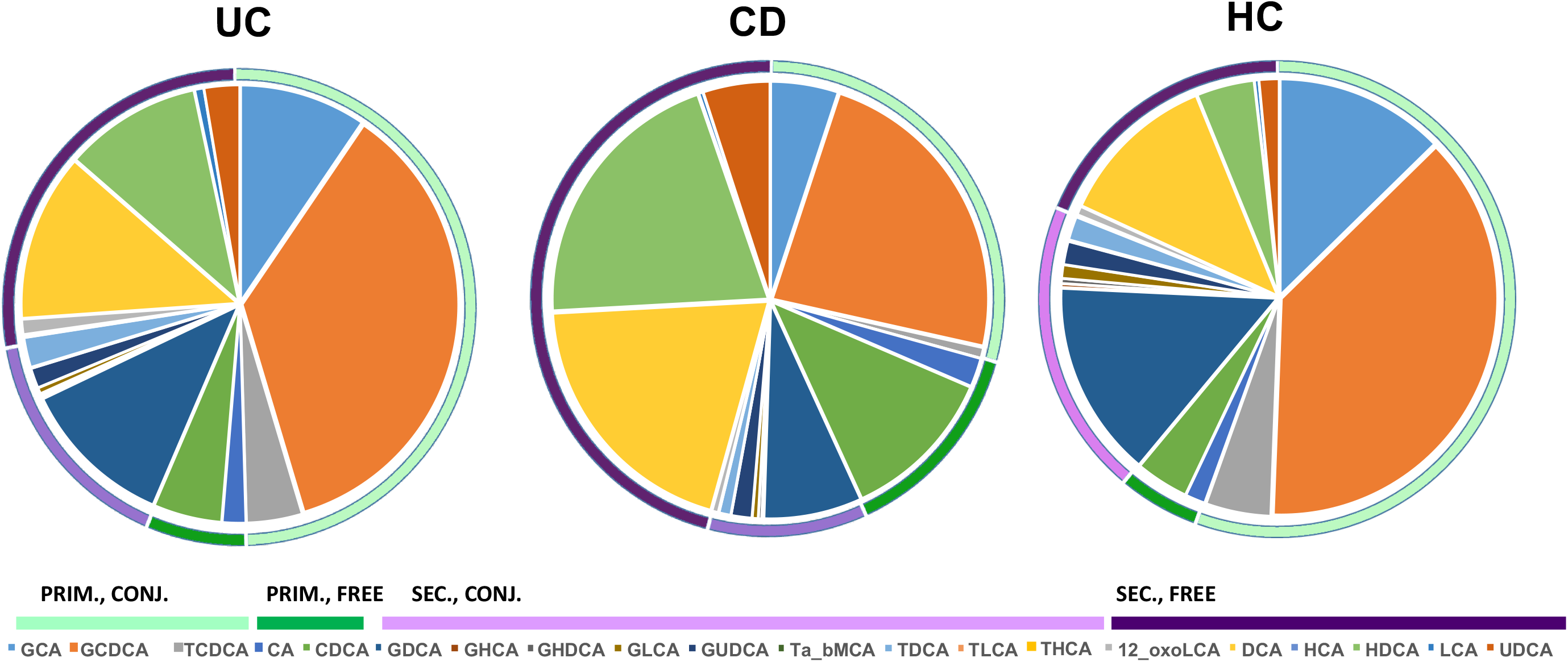
The distribution of individual BAs in UC, CD and HC, grouped as primary conjugated (PRIM., CONJ.), primary free (PRIM., FREE), secondary conjugated (SEC., CONJ.) and secondary free (SEC., FREE).

Comparisons of BAs levels were further stratified for disease location in CD according to the Montreal classification (Table 4). Ileal involvement (L1 and L3) was associated with a prominent decrease of ∑TBA (*p*=0.01) and the glycine-conjugated BA glycohyocholic acid (GHCA, *p*=0.02). An increase was observed for both of the free primary BAs (CDCA, *p*=0.008 and CA, *p*=0.04), the secondary BAs hyodeoxycholic acid (HDCA, *p*=0.004) and ursodeoxycholic acid (UDCA, *p*=0.004) in patients with ileal involvement (Table 4). The low number of non-resected patients with ileal involvement (n=3) did not allow us to examine the influence of a previous resection among patients with late-onset CD.

**Table 4.**
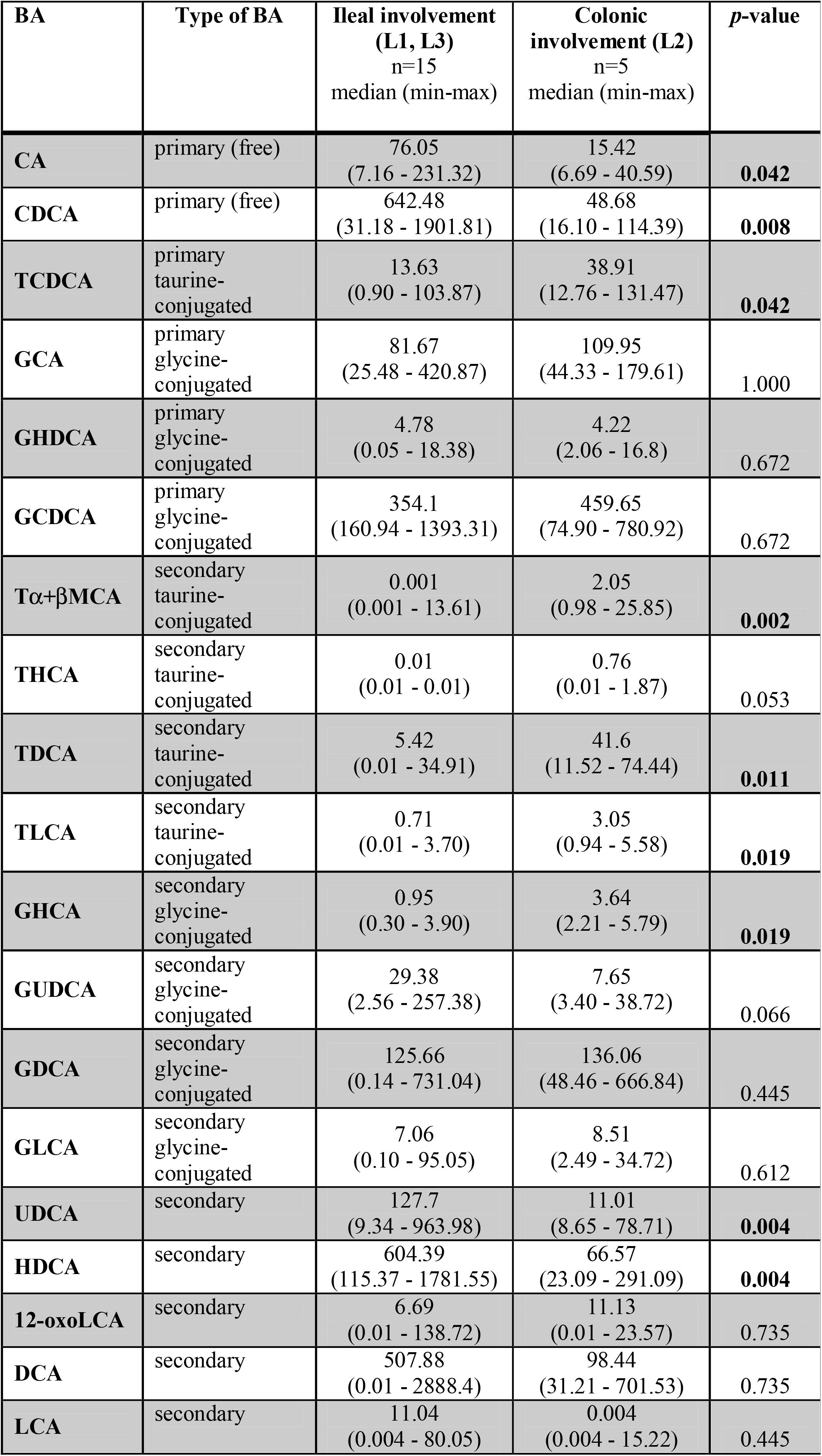
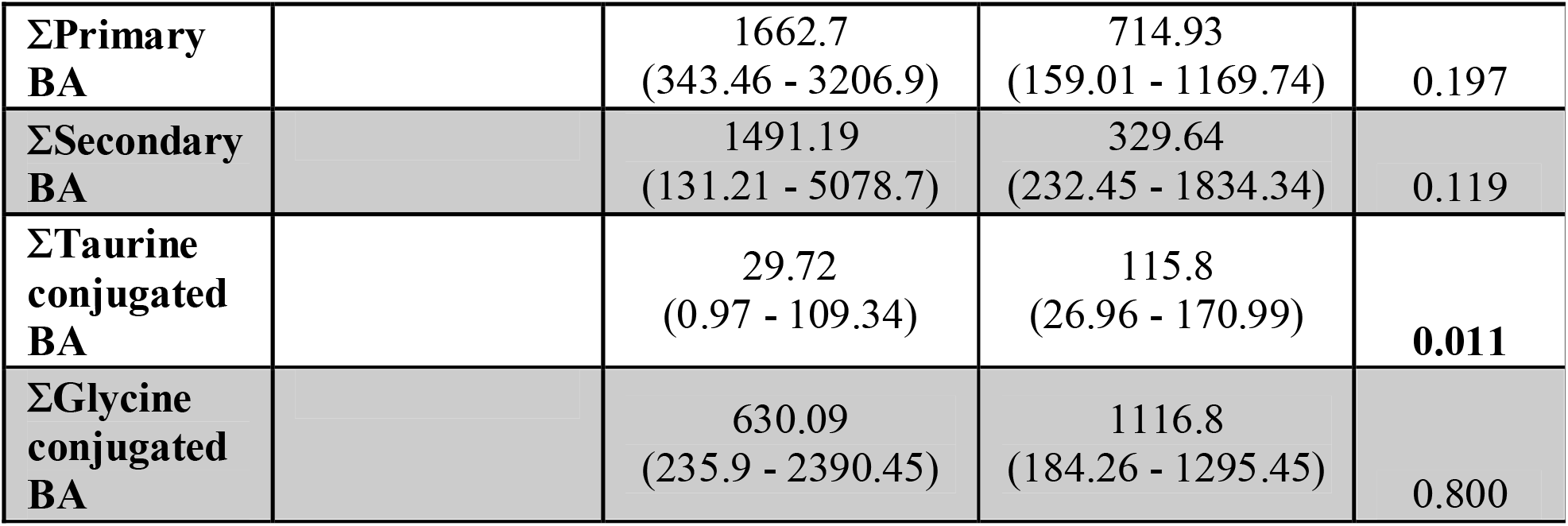
Differences in bile acid (BA) concentrations (ng/mL) in relation to Crohn’s disease location, according to Montreal classification. Significantly changing BA (*p*<0.05) are marked in bold.

| BA | Type of BA | Ileal involvement<br>(L1, L3)<br>n=15<br>median (min-max) | Colonic<br>involvement (L2)<br>n=5<br>median (min-max) | *p*-value |
| --- | --- | --- | --- | --- |
| CA | primary (free) | 76.05<br>(7.16 - 231.32) | 15.42<br>(6.69 - 40.59) | **0.042** |
| CDCA | primary (free) | 642.48<br>(31.18 - 1901.81) | 48.68<br>(16.10 - 114.39) | **0.008** |
| TCDCA | primary<br>taurine-conjugated | 13.63<br>(0.90 - 103.87) | 38.91<br>(12.76 - 131.47) | **0.042** |
| GCA | primary<br>glycine-conjugated | 81.67<br>(25.48 - 420.87) | 109.95<br>(44.33 - 179.61) | 1.000 |
| GHDCA | primary<br>glycine-conjugated | 4.78<br>(0.05 - 18.38) | 4.22<br>(2.06 - 16.8) | 0.672 |
| GCDCA | primary<br>glycine-conjugated | 354.1<br>(160.94 - 1393.31) | 459.65<br>(74.90 - 780.92) | 0.672 |
| Tα+βMCA | secondary<br>taurine-conjugated | 0.001<br>(0.001 - 13.61) | 2.05<br>(0.98 - 25.85) | **0.002** |
| THCA | secondary<br>taurine-conjugated | 0.01<br>(0.01 - 0.01) | 0.76<br>(0.01 - 1.87) | 0.053 |
| TDCA | secondary<br>taurine-conjugated | 5.42<br>(0.01 - 34.91) | 41.6<br>(11.52 - 74.44) | **0.011** |
| TLCA | secondary<br>taurine-conjugated | 0.71<br>(0.01 - 3.70) | 3.05<br>(0.94 - 5.58) | **0.019** |
| GHCA | secondary<br>glycine-conjugated | 0.95<br>(0.30 - 3.90) | 3.64<br>(2.21 - 5.79) | **0.019** |
| GUDCA | secondary<br>glycine-conjugated | 29.38<br>(2.56 - 257.38) | 7.65<br>(3.40 - 38.72) | 0.066 |
| GDCA | secondary<br>glycine-conjugated | 125.66<br>(0.14 - 731.04) | 136.06<br>(48.46 - 666.84) | 0.445 |
| GLCA | secondary<br>glycine-conjugated | 7.06<br>(0.10 - 95.05) | 8.51<br>(2.49 - 34.72) | 0.612 |
| UDCA | secondary | 127.7<br>(9.34 - 963.98) | 11.01<br>(8.65 - 78.71) | **0.004** |
| HDCA | secondary | 604.39<br>(115.37 - 1781.55) | 66.57<br>(23.09 - 291.09) | **0.004** |
| 12-oxoLCA | secondary | 6.69<br>(0.01 - 138.72) | 11.13<br>(0.01 - 23.57) | 0.735 |
| DCA | secondary | 507.88<br>(0.01 - 2888.4) | 98.44<br>(31.21 - 701.53) | 0.735 |
| LCA | secondary | 11.04<br>(0.004 - 80.05) | 0.004<br>(0.004 - 15.22) | 0.445 |
| **ΣPrimary**<br>**BA** |  | 1662.7<br>(343.46 - 3206.9) | 714.93<br>(159.01 - 1169.74) | 0.197 |
| **ΣSecondary**<br>**BA** |  | 1491.19<br>(131.21 - 5078.7) | 329.64<br>(232.45 - 1834.34) | 0.119 |
| **ΣTaurine**<br>**conjugated**<br>**BA** |  | 29.72<br>(0.97 - 109.34) | 115.8<br>(26.96 - 170.99) | **0.011** |
| **ΣGlycine**<br>**conjugated**<br>**BA** |  | 630.09<br>(235.9 - 2390.45) | 1116.8<br>(184.26 - 1295.45) | 0.800 |

### Correlations of PFAS and BAs

The total level of PFAS, (∑PFAS) was found to correlate positively with taurohyocholic acid, taurochenodeoxycholic acid (TCDCA) and ∑TBA among patients with late-onset UC (Figure 4). In addition, PFOS correlated positively with taurolitocholic acid (TLCA), while PFUnDA correlated positively with taurine-conjugates of α- and β-muricholic acid (Tα+βMCA) among these patients. On the contrary, PFDA showed a negative correlation with CA and TLCA. In CD, the total level of PFAS showed a positive correlation with taurochenodeoxycholic acid (TCDCA), GHCA and ∑TBA. PFNA and PFDA also correlated positively with Tα+βMCA. In HCs the correlation pattern was different compared to UC and CD, where ∑PFAS showed a negative correlation to UDCA and positive correlation to TLCA.

**Figure 4.**
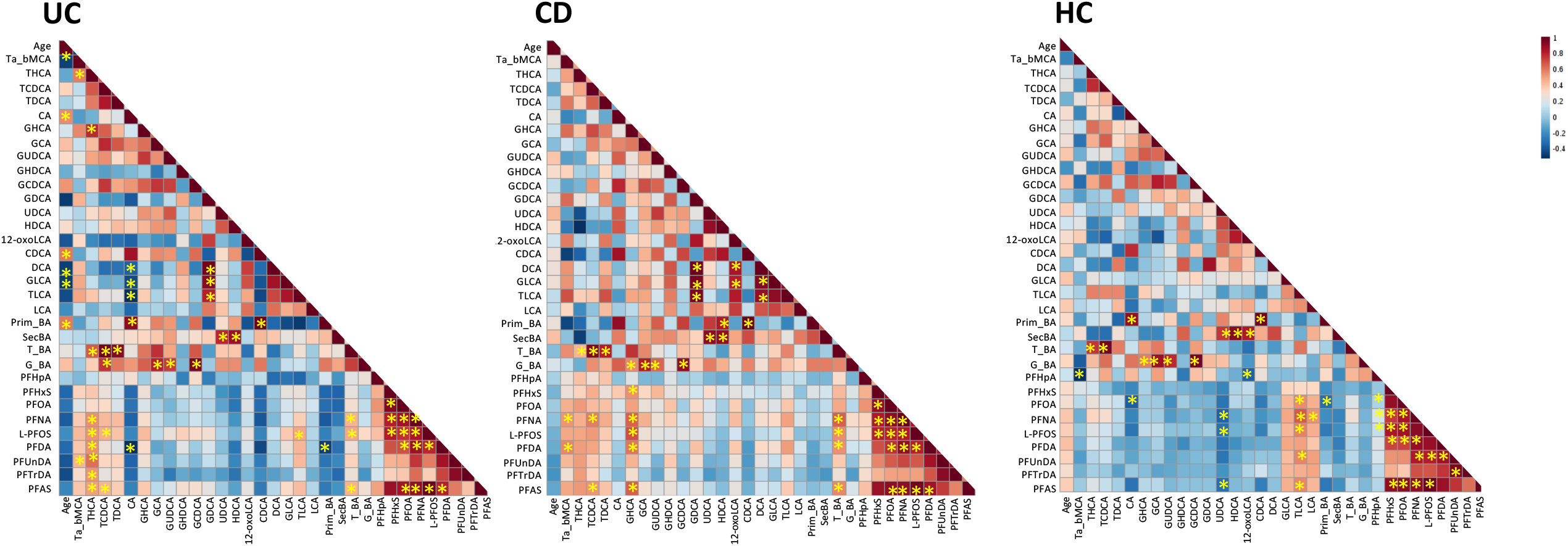
Spearman correlations between PFAS and BAs in A) UC B) CD and C) HC. Significant correlations (*p*<0.05) marked with a star (*).

## Discussion

Here we investigated the impact of PFAS on the risk of late-onset IBD, in relation to the intestinal barrier function and BA metabolism. Our findings demonstrate that serum PFAS levels, particularly PFOS and PFOA, are increased among late-onset UC patients. Interestingly, our findings were also associated with changes in the BA levels. Our hypothesis of a potential role for PFAS in IBD was further supported by our *ex vivo* study of mouse tissue, where exposure to PFOA resulted in increased ileal and colonic permeability and an enhanced colonic secretory response to CCh. Taken together, these findings may indicate that exposure to PFAS contribute to UC by disrupting the intestinal barrier or indirectly by interfering with the BA metabolism and thereby alter the intestinal barrier function. PFAS have been associated with various adverse health effects, including cancer, liver damage, decreased fertility, and increased risk of chronic inflammatory diseases [23]. Contamination of drinking water with high levels of PFOA, one of the most common PFAS, has also been associated with an increased incidence of UC in the state of Ohio, U.S. [7]. A recent study further showed that mice dosed with PFOA during 10 days displayed an altered expression of tight junction genes in the ileum compared to the colon [8]. These observations indicate that PFAS may play a role in the pathophysiology of IBD and potentially act as an environmental trigger. In the current study, we observed higher PFAS levels among patients with late-onset UC in comparison to HC and CD. Particularly, PFOS and PFOA were at higher levels among patients with UC. This finding is in line with a previous report showing higher levels of PFOA among patients with UC (n=116) [24]. However, a recent population study in Faroe island failed to show an increased PFAS level in 32 patients with UC and 5 patients with[25] CD (ref). This descripancy is most likely due to power issues or differences in the geographical exposure to PFAS as outlined by *Hammer T et al* [25].

Disruption of the intestinal barrier is believed to represent a key disease-mechanism in IBD [26,27]. Given the observed higher levels of PFOA in UC, we investigated the impact of *ex vivo* exposure of PFOA on ileal and colonic barrier function in mice [7,24]. Exposure to PFOA (10 and 100μM) induced an increased colonic and ileal permeability (G) as well as an increased ion transport across the colonic barrier (Isc) in response to stimulation with CCh. Taken together these data indicate that PFOA have the ability to not only affect the intestinal barrier directly but also decrease the resilience of the colonic barrier. To further support these findings we did observe a significantly increased ileal macromolecular passage after exposure to 10μM PFOA, but not after exposure to100μM PFOA, compared to vechile. Plausible explanation why we observed significant changes only at the lower exposure level is potentially due to the nonmonotonic response that has been reported in PFAS exposure[28,29]. A non-significant trend towards an increased macromolecular passage was observed across the colonic mucosa. The absence of a significant observation in colon could be due to lack of power. However, previous findings show that mice dosed with PFOA displayed more profound alterations in the ileal expression of genes encoding the tight junction proteins compared to colonic expression [8]. Hence, potential differences in the effect of PFOA on ileal and colonic mucosa will need to be addressed in future studies. Exposure to DMSO (vehicle) did result in an increased colonic permeability, particularly at 60 min. This is in line with previous findings showing that DMSO does have an effect on the intestinal epithelium and is therefore important to control for [30,31]. Notably, exposure to 100μM PFOA for 60 min did induce a significantly increased colonic permeability compared to DMSO alone (p<0.05, data not shown). A recent study, using zonulin as surrogate marker of intestinal permeability stated that there was no significant correlation between exposure to PFAS and zonulin levels among 179 patients/individuals [32], in contrast with our findings. However, the use of zonulin as a potential marker of intestinal permeability have recently been questioned ([33,34]. Currently, the Ussing Chamber is the most advanced methodology for investigation of the intestinal barrier function where both the electrophysiology and macropmolecular passage can be monitored, however it is important to acknowledge that our results are based on experiments in mice.

Plausible explanation of the changes in the intestinal permeability observed in our study could be linked with the PFOA-induced inhibition of Hepatocyte nuclear factor 4α (HNF4α) [35]), which known to be involved in the formation of tight junctions in the intestinal epithelium. Interestingly, HNF4α mRNA expression is significantly decreased in IBD patients [36] and it also plays a central role in the regulation of BA metabolism in the liver, where it is involved in both the synthesis and the conjugation of primary BAs [37]. Indeed, another potential mechanism via which PFAS exposure may contribute to a dysfunctional intestinal barrier and the pathophysiology of IBD is by affecting the composition of the BA pool [9,38,39]. *In vitro* studies have previously shown that direct exposure of PFAS to a hepatocyte cell line induces strong alterations in the intra- and extracellular BA content with a dose-dependent decrease of intracellular BAs occurring in response to PFAS. In the present paper, we identify significant differences in the circulating levels of BAs in UC and CD as compared to HC. An altered profile of circulating BAs in IBD has been reported previously [11,40,41]. Our results demonstrate that the total level of primary BAs is significantly decreased in UC and CD compared to HC. However, only patients with CD showed increased levels of the free primary CA and CDCA. Previous reports indicate that CDCA, even at low physiological levels (100 μM), has the potential to induce an increased mucosal permeability and bacterial uptake in human colonic tissue [13,42]. Moreover, the decrease in the ratio of CA/CDCA in CD, suggests that the synthesis of BAs is occurring *via* the alternative BA synthesis route (which usually accounts for only 10% of BA synthesis) and not *via* the classical pathway in the liver. This, in turn, suggests that CD is associated with a deficiency in the synthesis of BAs via the classical pathway.

The link between PFAS exposure and the composition of the BA pool is supported by the observed correlations between PFAS levels and the altered BA profile. In accordance with previous literature showing that PFAS suppress the biosynthesis of BAs, we observed negative correlations between the total exposure to PFAS and total levels of primary and secondary BAs [9,38] among HC, with only the secondary BA litocholic acid and its conjugates (GLCA, TLCA) showing a positive correlation. In addition, IBD patients showed a different profile with positive correlations between total ∑PFAS and ΣTBA. This indicates a possible alteration of the deconjugation in the gut, alternatively an increased absorption of taurine conjugates from the intestine. However, the changes observed in BA content in regard to CD, should be interpreted with caution, as most patients had ileal or ileocolonic CD. Both inflammation and resection of the terminal ileum are known to reduce the reabsorption of BA, and in accordance with previous studies [41,43], ileal CD was associated with an increase of primary BAs (CA and CDCA), an increase of secondary BAs (HDCA and UDCA), and a decrease in ΣTBA. The extent to which these alterations occur due to ongoing inflammation, previous resections, pronounced alterations in the composition and volatility of the gut microbiome or a combination of these could not be examined due to the small sample size [44,45].

The current study has some limitations. First of all, the number of individuals in the study was low, and results may have been influenced by previous or ongoing treatments, even though only one CD patient was prescribed BA sequestrants. Secondly, the HC group was slightly younger (mean age: 59) than the patients with IBD, and their serum samples were obtained a few years later. However, possible confounding effects of age and year of sampling were accounted for in our statistical analysis and should not have any major impact on the results. Nevertheless, future studies should validate these results in a larger cohort, where samples from cases and controls are obtained within an identical period. Additionally, we were not able to relate our findings to the composition of gut microbiota, since stool samples were not available. Future studies should investigate this as recent evidence indicate that PFAS can directly affect the gut microbiota composition. In the present study, we selectively included patients with late-onset IBD, based on the hypothesis that exposure to environmental risk factors is more important among these patients. The extent to which our findings can be generalised to patients with younger onset IBD is unknown. There is also a need to validate the observed effects on the intestinal barrier in response to exposure to PFOA in functional studies of mucosal samples from patients with IBD. In conclusion, our study demonstrates that total levels of PFAS are significantly increased in patients with late-onset UC compared to HC and patients with CD and correlate with disturbances in the BA pool. Moreover, we propose that a possible mechanism through which PFAS contribute to the pathophysiology of UC is *via* disrupting the intestinal barrier or indirectly by interfering with the BAs metabolism and that this subsequently contribute to an altered intestinal barrier function.

## Data Availability

The datasets generated during the current study are available from the corresponding author on reasonable request.

## Acknowledgement

We thank Dr Ciara Keogh and laboratory technician Jessica Sladek for skillful technical assistance in the Ussing laboratory, Davis, USA. This work was supported by funding from the Faculty of Medicine and Health, Örebro University [grant number: ORU2018/04457 to I.S], Bo Rydin foundation [grant number: F0514 to I.S], Örebro Hospital Research Foundation [Grant number OLL-790011 to J.H] Swedish research council [grant number 2016-05176 to T.H] and Formas [grant number. 2019-00869 to T.H and M.O].

