## Supplemental table 1 for "Perfluoroalkyl Substances are increased in patients with Late-Onset Ulcerative colitis and induce Intestinal Barrier defects *ex vivo* in Murine Intestinal tissue"

**Table A1**. Serum concentrations (ng/ml) of bile acids (BAs) and perfluoroalkyl substances (PFAS) in ulcerative colitis (UC), Crohn’s disease (CD) and healthy controls (HC). Table shows median concentrations with min/max.

|  |  | UC  ng/mL  median (min/max) | CD  ng/mL  median (min/max) | HC  ng/mL  median (min/max) |
| --- | --- | --- | --- | --- |
| *Bile acids (BAs)* |  |  |  |  |
| CA,  Cholic acid | primary (free) | 24.98  (4.02-515.49) | 37.63  (6.69-231.32) | 30.21  (5.14-537.29) |
| CDCA,  Chenodeoxycholic acid | primary (free) | 73.07  (4.35-2700.77) | 203.45  (16.10-1901.81) | 80.11  (9.87-606.25) |
| TCDCA,  Taurochenodeoxycholic acid | primary  taurine-conjugated | 59.29  (4.10-445.43) | 13.92  (0.90-131.47) | 97.92  11.39-371.25 |
| GCA, Glycocholic acid | primary  glycine-conjugated | 135.09  (28.68-648.78) | 87.81  (25.48-420.87) | 254.40  (64.07-1039.72) |
| GHDCA,  Glycohyodeoxycholic acid | primary  glycine-conjugated | 2.23  (0.03-30.71) | 4.50  (0.03-18.38) | 7.60  (0.03-46.41) |
| GCDCA,  Glycochenodeoxycholic acid | primary  glycine-conjugated | 511.02  (112.04-4373.75) | 406.88  (74.90-1393.31) | 761.93  (90.22-2113.96) |
| Tα+βMCA, Taurine-conjugates of  α- and β- muricholic acid | secondary  taurine-conjugated | 0.96  (0.00-7.91) | 0.20  (0.00-25.85) | 1.97  (0.00-17.94) |
| THCA,  Taurohyocholic acid | secondary  taurine-conjugated | 0.21  (0.01-8.68) | 0.01  (0.01-1.87) | 1.60  0.01-7.71 |
| TDCA,  Taurodeoxycholic acid | secondary  taurine-conjugated | 31.90  (2.04-294.87) | 15.36  (0.00-74.44) | 35.69  2.37-113.93 |
| TLCA,  Taurolitocholic acid | secondary  taurine-conjugated | 2.92  (0.05-37.08) | 0.93  (0.01-5.58) | 3.19  (1.09-12.60) |
| GHCA,  Glycohyocholic acid | secondary  glycine-conjugated | 2.85  (0.80-20.36) | 2.04  (0.30-5.79) | 5.66  (2.50-15.89) |
| GUDCA,  Glycoursodeoxycholic acid | secondary  glycine-conjugated | 21.46  (4.94-312.86) | 26.99  (2.56-257.38) | 34.94  (4.49-264.67) |
| GDCA,  Glycodeoxycholic acid | secondary  glycine-conjugated | 163.43  (0.10-974.91) | 127.39  (0.10-731.04) | 295.31  (0.10-2154.80) |
| GLCA,  Glycolitocholic acid | secondary  glycine-conjugated | 6.55  (0.10-46.22) | 7.78  (0.10-95.05) | 20.32  (4.25-141.00) |
| UDCA,  Ursodeoxycholic acid | secondary (free) | 37.75  (2.74-175.88) | 86.92  (8.65-963.98) | 30.12  (16.76-194.31) |
| HDCA,  Hyodeoxycholic acid | secondary (free) | 146.02  (11.78-990.55) | 356.95  (23.09-1781.55) | 86.99  (8.48-498.08) |
| 12-oxoLCA,  12-oxo-lithocholic acid | secondary (free) | 16.16  (0.01-86.13) | 8.34  (0.01-138.72) | 13.26  (2.45-34.82) |
| DCA,  Deoxycholic acid | secondary (free) | 178.19  (0.01-1255.20) | 344.10  (0.01-2888.40) | 240.82  (0.01-737.10) |
| LCA,  Lithocholic acid | secondary (free) | 9.75  (0.00-24.50) | 5.40  (0.00-80.05) | 6.73  (0.00-21.96) |
| ΣPrimary BA | free and conjugated | 709.73  (194.72-5549.51) | 578.53  (138.69-1802.93) | 1186.44  (174.59-4062.22) |
| ΣSecondary BA | free and conjugated | 644.78  (111.04-2402.54) | 1381.47  (131.94-5082.87) | 868.72  (156.46-2405.24) |
| ΣTBA,  Taurine-conjugated BA | primary and secondary | 630.09  (7.35-743.20) | 39.65  (0.97-170.99) | 187.16  (18.69-452.24) |
| ΣGBA,  Glycine-conjugated BA | primary and secondary | 919.49  (215.84-6214.40) | 142.46  (184.26-2390.45) | 1462.26  (260.92-5412.31) |
| Per-and polyfluoroalkyl substances  *(PFAS):* |  |  |  |  |
| PFHpA,  Perfluoroheptanoic acid |  | 0.07  (0.04-3.79) | 0.05  (0.04-0.20) | 0.04  (0.04-0.14) |
| PFHxS,  Perfluorohexane sulfonate |  | 1.04  (0.17-5.80) | 0.98  (0.16-2.98) | 0.78  (0.07-6.18) |
| PFOA,  Perfluorooctanoic acid |  | 2.98  (0.61-14.84) | 2.06  (0.47-7.30) | 1.42  (0.15-3.48) |
| PFNA,  Perfluorononanoic acid |  | 0.89  (0.13-3.21) | 0.57  (0.11-1.71) | 0.76  (0.06-2.03) |
| PFOS,  Perfluorooctane sulfonate |  | 8.68  (1.87-111.87) | 4.62  (0.28-21.53) | 4.20  (0.44-16.69) |
| PFDA,  Perfluorodecanoic acid |  | 0.39  (0.13-1.20) | 0.20  (0.03-0.89) | 0.37  (0.04-0.84) |
| PFUnDA,  Perfluoroundecanoic acid |  | 0.28  (0.10-0.97) | 0.20  (0.08-1.06) | 0.39  (0.06-0.91) |
| PFTrDA,  Perfluorotridecanoic acid |  | 0.06  (0.02-0.29) | 0.06  (0.01-0.39) | 0.08  (0.01-0.20) |
| ΣPFAS,  Total PFAS-levels |  | 16.82  (3.81-141.71) | 9.99  (2.61-36.81) | 8.25  (1.71-29.23) |
